# The predictive role of pain catastrophising following genicular arterial embolisation for the treatment of mild and moderate knee osteoarthritis

**DOI:** 10.1101/2023.07.31.23290995

**Authors:** Richard Harrison, Tim V. Salomons, Sarah MacGill, Mark W. Little

**Affiliations:** Centre for Integrative Sciences and Neurodynamics, University of Reading, School of Psychology and Clinical Language Sciences, Reading, UK; University Department of Radiology, Royal Berkshire NHS Foundation Trust, Reading, UK; Pain Affect and Cognition Lab, Queen’s University, Department of Psychology, Kingston, ON, Canada

**Keywords:** Pain, Catastrophising, MRI, DLPFC, Embolisation, Osteoarthritis, Postsurgical Chronic Pain, Prediction, Surgical

## Abstract

Knee osteoarthritis (OA) is the most common form of OA and is not currently considered to be a curable disease. Specifically, mild-to-moderate knee OA that is resistant to conservative treatment, but does not warrant joint replacement, poses a significant clinical problem. Genicular arterial embolisation (GAE) is an interventional radiological technique designed to subvert neoangiogenesis within the joint, in turn reducing pain and improving function. Preliminary data has identified a subset of patients who do not respond, despite a technically successful procedure. We therefore investigated individual differences in pain and pain perception to identify predictive pre-surgical markers for clinical outcomes. Specifically, we investigated pain catastrophising (PC) and its neural correlates using resting-state functional magnetic resonance imaging (rs-fMRI). Thirty patients participated in a presurgical assessment battery during which they completed psychometric profiling and quantitative sensory testing. A subset of seventeen patients also completed an rs-fMRI session. Patients then recorded post-surgical outcomes at 6-weeks, 3-months, 12-months and 24-months. The dorsolateral prefrontal cortex (DLPFC) served as a seed for whole-brain voxel-wise connectivity with pain catastrophising scores entered as a regressor in group analysis. Pain catastrophising was associated with a myriad of aversive psychological/lifestyle variables at baseline, as well as a predisposition for attending to pain. Surprisingly, high pain catastrophisers stood to gain the best improvements from GAE, with PC scores predicting the higher reductions in pain across all time-points. Seed-based whole-brain connectivity revealed that PCS was associated with higher connectivity between the DLPFC and areas of the brain associated with pain processing, suggesting more frequent engagement of top-down modulatory processes when experiencing pain. These results are an early step towards understanding outcomes from novel interventional treatments for mild-to-moderate knee OA. Data suggests that improvements in pain and function via GAE could help high catastrophisers manage their pain, and in turn, the negative associations with pain that were identified at baseline.

## Introduction

It is estimated across a lifespan that 47% of women and 40% of men will develop symptomatic knee osteoarthritis[39]. Osteoarthritis (OA) is the most common form of arthritis[31], which in turn is a leading cause of disability globally[22]. At present, OA is not considered to be a curable disease, in part, because the pathophysiological mechanism is not yet comprehensively understood[36]. The goal of OA treatment is to slow the progression and alleviate the symptoms of the disease.

Traditionally, initial treatment consists of conservative options such as physiotherapy, orthotics and pharmacology. It is only when all conversative options are exhausted, and the pathological indicators reach a threshold of severity, that surgical options are considered[13,38]. Given the central role of psychosocial factors in the maintenance of chronic knee pain[37], a prolonged period of poor treatment response can have serious implications for the entrenchment of chronic pain. Mild-to-moderate OA resistant to nonsurgical options, yet not severe enough to warrant join replacement surgery, poses a significant management problem. Genicular artery embolisation (GAE) is a novel interventional radiological technique that is easier to deliver and less invasive which is designed to subvert neoangiogenesis within the joint, hypothesised to contribute to structural damage and pain in knee OA [30,34,40].

While promising, preliminary data for GAE indicates a subset of patients who do not respond to treatment, despite a technically successful procedure [34]. These findings are also mirrored within more invasive surgical options such as total knee replacement (TKR), wherein 6-30% of patients continue to experience chronic pain after the procedure[2,7,10]. It is clear that a strictly pathophysiological approach to the assessment of pain is ineffective[8] and that to understand the interpatient variability in outcomes we must complement these assessments with psychosocial insight. Moreover, complementing these assessments with predictive perspectives may facilitate earlier intervention, to help disrupt the negative progression of the chronic pain cycle.

Pain catastrophising (PC) is a cognitive-affective bias characterised by a negative interpretation of the consequences of pain[42,52]. Catastrophising is regularly quantified using the Pain Catastrophising Scale[51], and comprises of three main elements; rumination, magnification and helplessness. PC has been associated with lower pain thresholds[16] and poor longitudinal surgical response to a range of conditions[28,41], including knee osteoarthritis[10,15,43]. Alongside it’s predictive capabilities, it has been shown to be modifiable by psychological intervention[17,18,33], underlining the potential value for presurgical identification to improve outcomes.

PC is frequently characterised as a cognitive attentional bias to the processing of nociceptive stimuli. Evidence suggests catastrophising is not directly associated with the sensory-discriminative dimensions of pain, but instead the modulation and processing of the unpleasantness of pain[46]. Attentional regulation has been found to also play a key role in catastrophising, with high catastrophisers often struggling to disengage attention from pain[12]. As such, neuroscientific investigation focuses on regions such as the dorsolateral prefrontal (DLPFC), anterior cingulate (ACC), insula and medial prefrontal cortices. These cortical regions are associated with the emotional, and attentional modulation of pain, as well as pain salience, vigilance and awareness. Specifically, the dlPFC is thought to play a role in top-down modulation, underlying the facilitatory influence of PC on pain[19,24,35,47,48]. Therefore, while PC is not directly associated with the sensory-discriminative response to pain, the processing and suppression of this sensory information may underlie the maladaptive influence of catastrophising[42,46]. Furthermore, the manner in which individuals attend to pain involves an interaction between top-down and bottom-up influences and is likely to facilitate catastrophisation[12].

In the current study, we examined pre-surgical characteristics of patients and tested the hypothesis that clinical outcomes following GAE in patients with mild to moderate knee OA can be predicted by presurgical pain catastrophising levels. Secondly, we investigated the influence of pain catastrophising on Intrinsic Attention to Pain (IAP), specifically, whether catastrophisers have a higher tendency to attend towards pain. Lastly, to better understand intrinsic neural mechanisms, we used resting-state fMRI (rs-fMRI) to examine dlPFC-to-whole brain neural connectivity at rest. We hypothesised that the neural mechanism underlying PC would be associated with variable connectivity of the dlPFC, as a key region in pain modulation and the suppression of pain intensity[35], and regions of the brain associated with sensory processing of nociceptive signals, such as the motor and somatosensory cortices[54,58].

## Methods

### Sample

Thirty-five patients with a diagnosis of mild to moderate knee OA volunteered for a collaborative research study between the University of Reading and the Royal Berkshire NHS Foundation Trust (RBFT) (The GENESIS Study IRAS: 237676, CPMS: 37741). All patients consented to procedures approved by the Health Research Authority (HRA), the NHS London Bromley Research Committee and the University of Reading Research Ethics Committee (UREC). Two patients declined to participate in the study after the procedure was completed, two patients decided to pursue referrals for knee replacement and another patient discontinued their participation due to cumulative delays caused by Covid-19. Within this sample, a subset of twenty patients agreed to take part in a neuroimaging session, of which three patients did not complete the MRI scan due to claustrophobia. This left a final behavioural sample of thirty patients (M_age_= 61.7, s.d= 11.1; 15 females) and a final neuroimaging sample of seventeen patients (M_age_= 58.7, s.d=9.5; 10 females).

The inclusion criteria for the study were a minimum age of 45, a diagnosis of mild-to-moderate knee OA and knee pain for at least 6 months, which was resistant to conversative treatment methods. Patients were excluded if they had infectious or rheumatoid arthritis, severe knee OA, renal impairment, bleeding diathesis, irreversible coagulopathy or previous knee arthroplasty. Patients were also required to have acceptable comprehension of English and have no MRI contraindications.

## Materials

### Thermal Stimulation

Noxious heat stimuli were generated using a MEDOC Pathway system (Ramat-Yishai, Israel), with a 30×30cm Peltier thermode. The thermode was securely attached to the underside of the lower right arm, with the patient’s arm resting on their upper thigh for comfort and to keep the thermode stable.

### Presurgical Questionnaires

Prior to the embolisation, patients completed a series of questionnaires designed to quantify psychological aspects that have been previously associated with poor surgical outcomes[14,32,43,57]. Pain catastrophising was quantified using the Pain Catastrophising Scale (PCS)[51], a 13-item measure scored using a 5-point Likert scale (0=not at all to 4= all the time). The PCS can be used as a unidimensional measure, or can be subdivided into three subscales; rumination, magnification or helplessness.

The other included measures were the Five Factor Mindfulness Questionnaire(FFMQ)[1], a 39-item measure used to quantify an individual’s intrinsic propensity to mindfulness. The State-Trait Anxiety Inventory (STAI)[50] and Becks Depression Inventory (BDI)[3] were used to quantify anxiety and depressive symptoms, respectively. Lastly, sleep quality was quantified using the Pittsburgh Sleep Quality Index (PSQI)[11].

### Outcome Questionnaire

Throughout participation in the study, patients completed two questionnaires to evaluate outcomes. The Knee Injury and Osteoarthritis Score (KOOS)[44] is a 42-item knee-specific measurement tool frequently used to evaluate treatment response for knee OA by assessing patients opinions about their knee. The KOOS is a multi-dimensional tool with 5 distinct subfactors; Pain, function in daily life, other symptoms, function in sports & recreation and quality of life. Scores on the KOOS are calculated as a percentage of total score achieved (0-1), with lower scores signifying worse outcome. The KOOS is the extension of another measure, the Western Ontario and McMaster Universities Arthritis Index (WOMAC)[5]. Items used to calculate a WOMAC score are contained within the KOOS, and represent a unidimensional measure for the impact of osteoarthritis pain[45], providing utility for the prediction of a univariate dependent variable. For this reason, the WOMAC was chosen as a primary outcome variable. Lastly, patients were asked to provide a 0-100 pain intensity rating specifically for their knee, using a numeric rating scale (NRS; 0: “No pain at all”;100: “The most intense pain imaginable”). All outcome measures were collected at baseline, and post-surgically at 6-weeks, 3-months, 12-months and 24-months.

### Design

The current study forms part of the GENESIS study, the interim analysis of which provides a full description of the GAE assessment protocol and procedure[34]. After the initial clinical assessment, patients attended a single-session assessment at the University of Reading. Patients firstly completed a sensory pain assessment. After this, patients either completed the presurgical questionnaires, or provided the questionnaires printed and completed from home within 7 days of the assessment. Lastly, patients then completed a neuroimaging session.

### Procedure

#### Sensory Pain Assessment

Firstly, pain thresholds were calibrated using a dual method approach. Both methods utilised an NRS, anchored with 0 as “no pain” and 10 as “most intense pain”. This dual-method approach utilised a method-of-limits and method-of-levels design, with the ultimate threshold being calculated as the average of these two tests. For a full description of the procedure, please refer to Harrison et al., 2018[23].

After the calculation of threshold, patients completed an intrinsic attention to pain (IAP) task, adapted from Kucyi et al., 2014[29]. An initial temperature calibration was completed with a dummy 20 second stimulus set at threshold+1°C. If patients provided a rating outside of 5-7/10, the stimulus temperature was raised or lowered by 0.5°C, and calibration was restarted. This process repeated until a rating between 5-7/10 was given. The IAP task was completed in silence and consisted of 10 consecutive 20-second stimuli with a 30-second ISI at a baseline temperature of 32°C, and ramp rate of 8°C. After each stimulus, a rating was provided using a different NRS, wherein patients rated to what extent they had been thinking about pain or something else (−2: Only something else, −1: Mostly something else, 1: Mostly pain, 2: Only pain). IAP score was calculated as an average of the 10 ratings, with a positive value indicating a proclivity for attention to pain.

#### fMRI Acquisition

Functional images were acquired using a Siemens MAGNETOM Prisma 3T scanner (Siemens, Erlangen, Germany), using a 64-channel head and neck coil. The protocol consisted of an initial localiser, followed by a resting-state scan, in which patients were instructed to lie still, and keep their eyes open. Functional data were acquired using a blood-oxygen level-dependent (BOLD) protocol with a T2*-weighted gradient echo planar imaging sequence (TR= 1000ms, TE= 30s, slice thickness= 2mm, FA= 90°, 256×256 matrix, voxel size= 2×2×2, FOV=256mm). To reduce the impact of field inhomogeneity, an initial 5 volumes were discarded, and subsequently 600 volumes were acquired, equally a total scan time of 10 minutes and 28 seconds. Following the resting-state, two field maps were collected, followed by a 5-minute T1-weighted inversion recovery fast gradient echo high-resolution anatomical scan (TR= 2300ms, TE= 2.29ms, slice thickness= 0.94mm, FA=8°, 256×256 matrix, voxel size= 0.9×0.9×0.9, FOV=240mm).

#### Behavioural Analysis

To evaluate the efficacy of the surgical procedure, data were inspected for normal distribution and subsequently a repeated-measures ANOVA was performed to test for a difference in mean pain ratings across four time-points (baseline/6-week/3-months/12-months). In the case of a significant mean effect, paired t-tests were then performed as post-hoc tests evaluating the differences in WOMAC ratings between baseline, and the three subsequent time-points (6-weeks, 3-months, 12-months, 24-months). To assess the relationship between presurgical baseline measures, Pearson’s correlations were performed for catastrophising (PCS), mindfulness (FFMQ), sleep quality (PSQI), anxiety (STAI) and depression (BDI) scores, as well as their relationship with baseline pain (WOMAC, KOOS_Pain and NRS). Additionally, to investigate the predictive capabilities of PCS, linear regressions were conducted with PCS as the independent variable, and WOMAC change from baseline as a dependent variable. For this analysis, change variables were coded by calculating the difference between 6-weeks/3-months/12-months/24-months and baseline, so that higher positive values represent higher pain intensity or higher detrimental impact of osteoarthritis. Regarding the sensory pain assessment, to evaluate how catastrophising may be associated with a tendency to attend to pain, correlations were conducted between PCS and IAP scores. The significance level was set to p<.05 for all analyses, which were completed using IBM SPSS Statistics 23 (IBM Corp. Version 23)

### fMRI Analysis

#### ROI Selection

For the purposes of preparing the ROI, a DLPFC mask was identified using the Harvard-Oxford cortical 100% probabilistic structural atlases. Due to no specific hypotheses regarding lateralised neural mechanisms within resting-state, a bilateral mask was then created using this method (Fig.1). The DLPFC was selected as a seed due to its prominent role in top-down modulation of pain[46,47], as well as its association with pain catastrophising[19,48].

**Figure 1.**
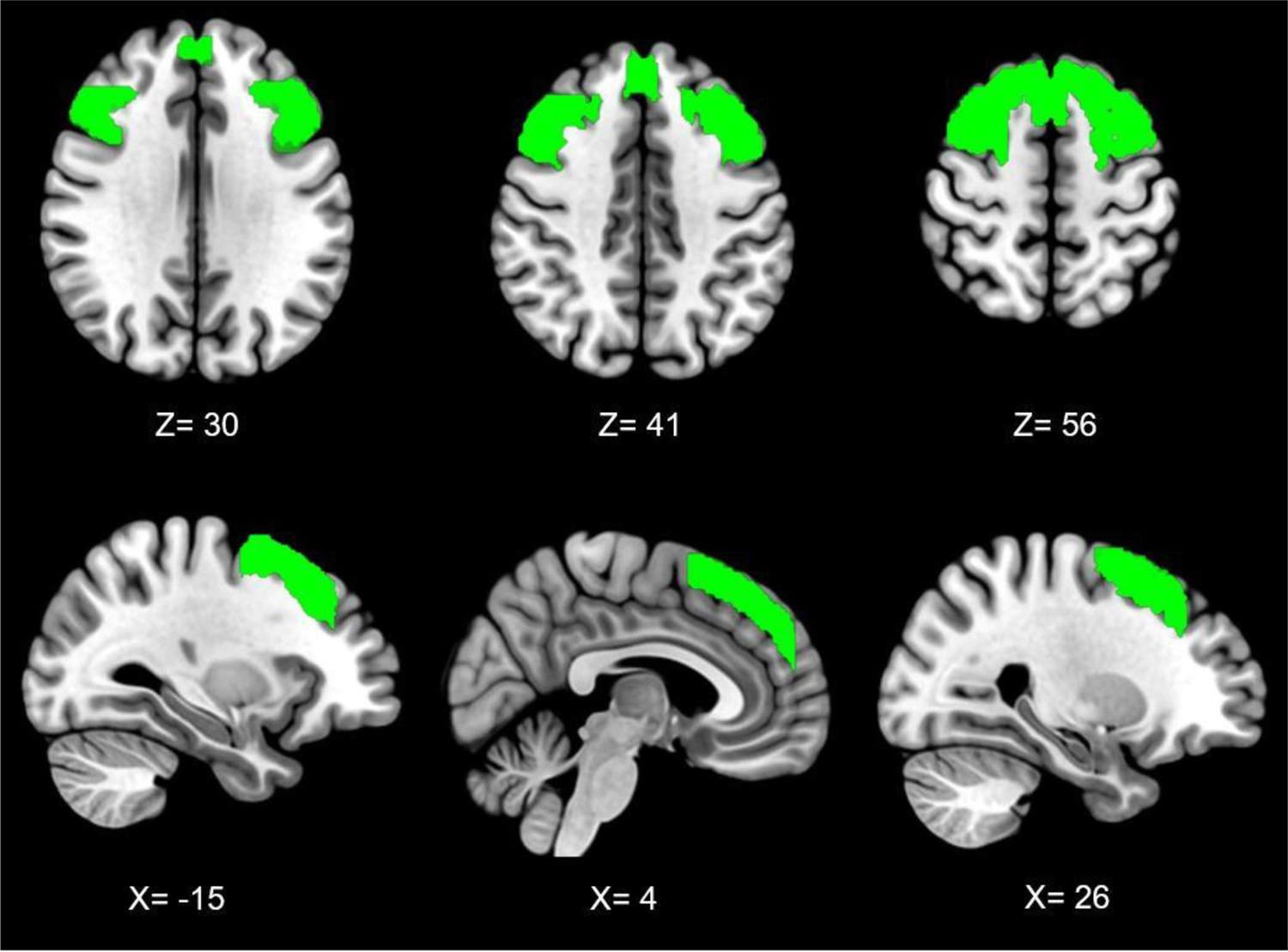
Bilateral dorsolateral prefrontal cortex seed across axial (top) and sagittal (bottom) planes, with co-ordinates shown in mm

**Figure 2.**
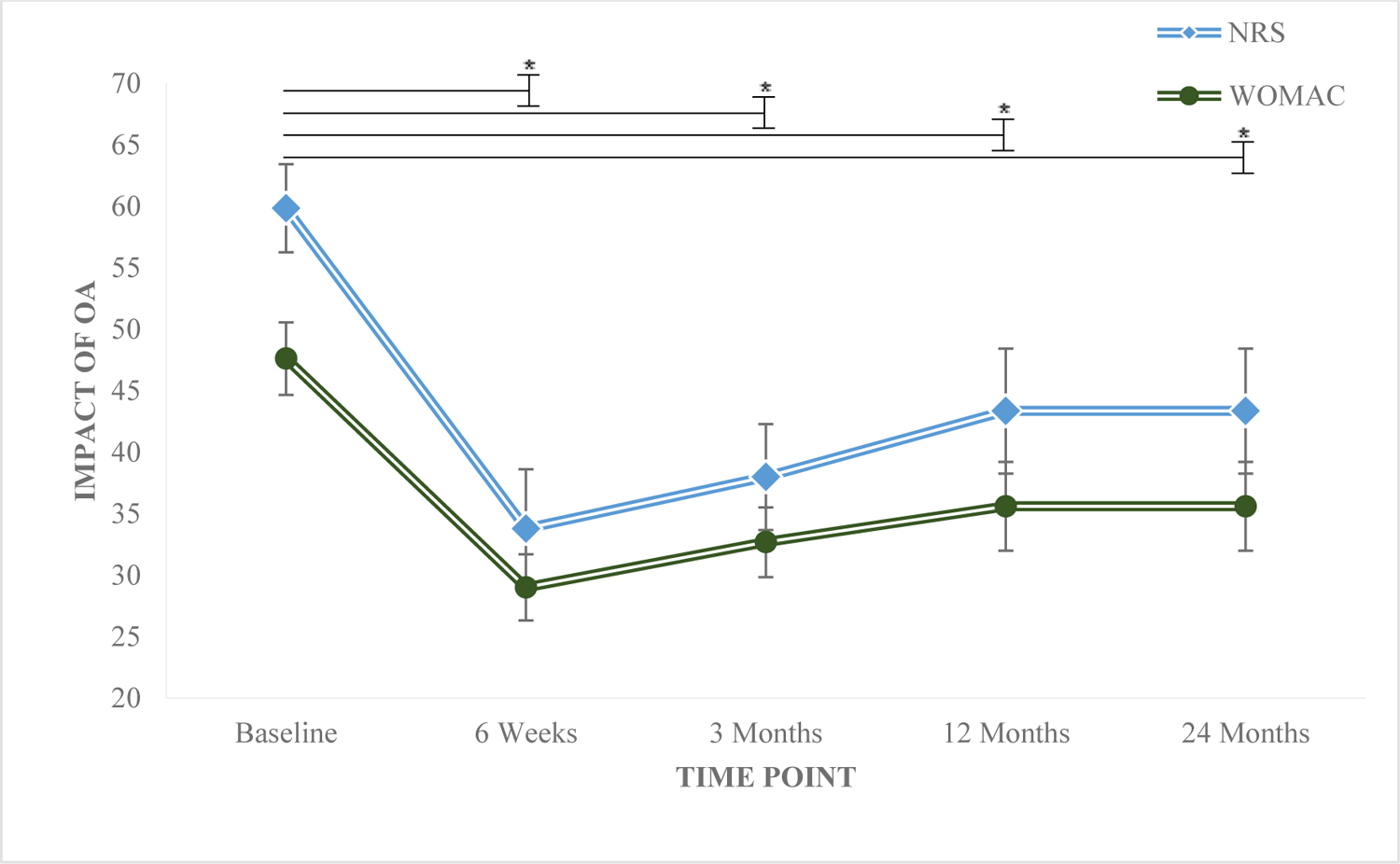
Longitudinal outcomes following successful completion of genicular arterial embolisation *(standard error bars)*. | Lower WOMAC and NRS values represent less severe impact of OA and lower pain, respectively. Stars denote significant improvements from baseline

#### Pre-processing

All analysis was performed using FMRIB’s Software Library Package (FSL 6.0[27]), following the Component Based Noise Correction Method (CompCor[4]). During acquisition, the first five volumes were discarded to facilitate signal equilibration. Correction for interleaved acquisition was applied and data were spatially smoothed using a 5mm full-width half-maximum (FWHM) gaussian kernel. The Brain Extraction Tool (BET[49]) was used for skull stripping. MCFLIRT[26] was used for the purposes of motion correction, and data were visually inspected to identify problems with registration, inadequate skull-stripping or uncorrected motion artifacts. To reduce the influence of non-neuronal activity, FAST[59] was then used to created segmented white matter (WM) and cerebrospinal fluid (CSF) masks, which were then thresholded to 0.99. Time-series were then extracted for each participant and added to the GLM as nuisance variables. Residuals from pre-processing and nuisance removal were then normalised and band-pass filtered (0.1/0.001Hz) to reduce the influence of high-frequency (i.e. cardiac/respiratory) and low-frequency (i.e. scanner drift) factors.

#### Resting-state Connectivity Analysis

The standardised DLPFC mask was registered to single-subject space, and a mean time series of all voxels within the ROI were extracted and added as an explanatory variable within a whole-brain functional connectivity analysis. Resulting contrast maps were then used as inputs within a higher-level analysis, alongside patient’s normalised pain catastrophising scores. The purpose of this analysis was to identify regions were connectivity to the DLPFC was associated with individual differences in PCS. Using FEATQuery, parameter estimates were extracted from resulting significant clusters for the purposes of the graphical representation of connectivity. Multiple comparisons corrections were applied using the Gaussian random field theory (Z<2.3;p<.05).

## Results

### Embolisation Pain Outcomes

At baseline, the mean NRS pain rating was 60.7/100 (s.d=19.4), which reduced to 33.8 (s.d=25.5) six-weeks post-surgery. Pain ratings remained lower than baseline at 3-months (M=37.9, s.d.=23.6), 12-months (M=43.3,s.d=26.4) and 24-months (M=39, s.d=26.8). Embolisation resulted in improvements in osteoarthritis outcomes, as quantified via WOMAC, from baseline to 6-weeks(t(28)=4.9,p<.001), 3-months(t(29)=3.9,p<.001), 12-months(t(26)=2.7,p=.013) and 24-months (t(19)=2.7,p=.013). This was supported by the same significant reductions in NRS and the pain subscale in the KOOS across all outcome points. This data, alongside that previously published by Little et al.,[34], indicates that embolisation is a suitable treatment for the management of mild/moderate osteoarthritis.

### Presurgical Baseline Psychometrics

Pain catastrophising scores were significantly correlated with all other baseline measures, with the exception of NRS. At baseline, pain catastrophising was associated with poor sleep quality (r(29)=.40,p=.03), low trait mindfulness (r(28)= −.45,p=.02), depression (r(29)=.41,p=.03), anxiety (r(29)= .53,p=.003) and higher pain via the KOOS (r(30)= −.45,p=.01) and WOMAC (r(30)=.39,p=.03), whereas the NRS was a non-significant trend (r(30)=.29,p=.12). PCS was also significantly correlated with IAP (r(30)=.53,p=.002), indicating that higher catastrophisers are more likely to attend to pain (Fig.3).

**Figure 3.**
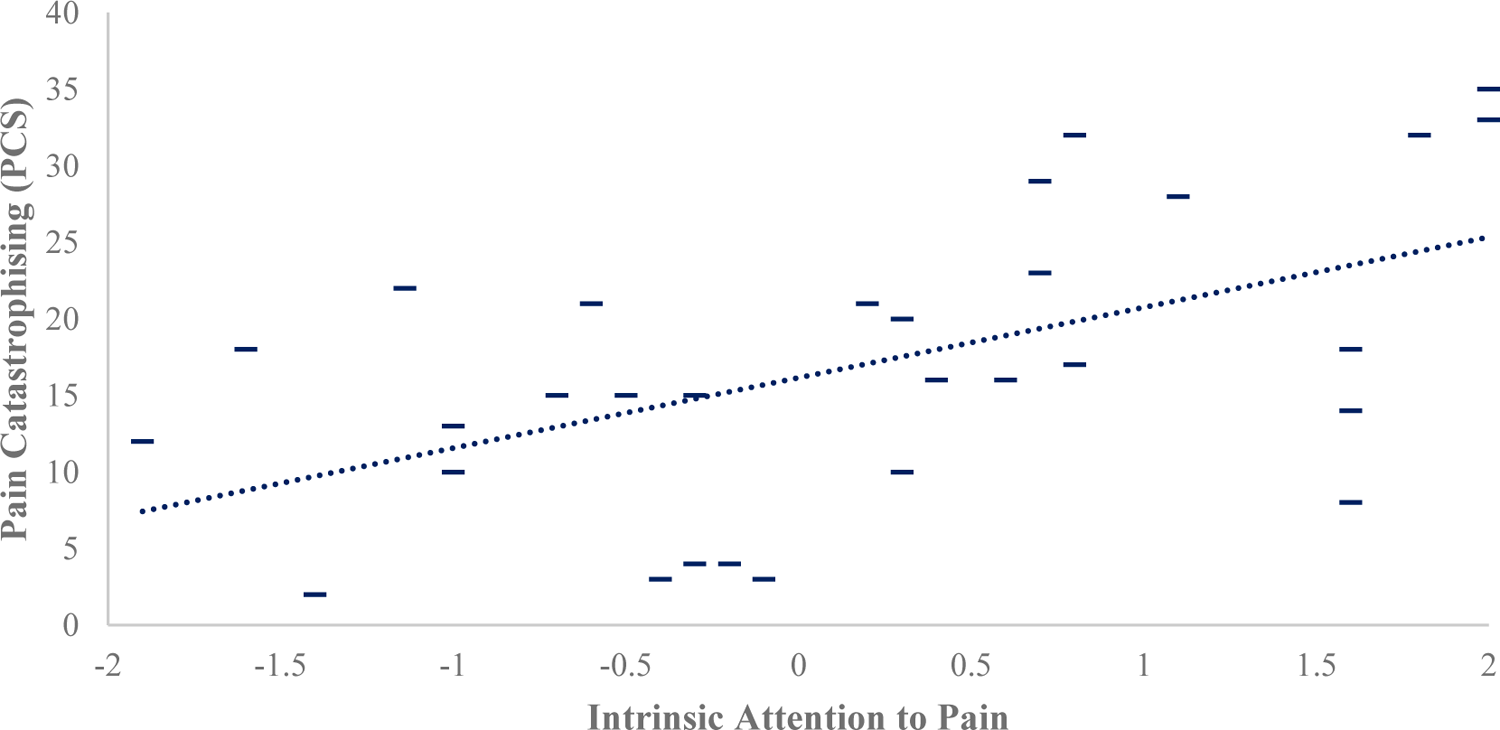
Association between pain catastrophising and intrinsic attention to pain. | Positive values on IAP represent attention to pain, negative values represent attention to something else.

**Table 1.** Association between presurgical baseline variables.

|  | PCS | FFMQ | PSQI | BDI | STAI | KOOS<sub>Pain</sub> | NRS | WOMAC |
| --- | --- | --- | --- | --- | --- | --- | --- | --- |
| PCS |  | -.45* | .40* | .41* | .53** | -.45* | .29 | .39* |
| FFMQ | -.45* |  | -.32 | -.62** | -.61** | .24 | -.13 | -.22 |
| PSQI | .40* | -.32 |  | .61** | .42* | -.41* | .40* | .40* |
| BDI | .41* | -.62** | .61** |  | .50** | -.27 | .25 | .37* |
| STAI | .53** | -.61** | .42* | .50** |  | -.22 | .20 | .17 |
| KOOS<sub>Pain</sub> | -.45* | .24 | -.41* | -.27 | -.22 |  | -.51** | -.87** |
| NRS | .29 | -.13 | .40* | .25 | .20 | -.51** |  | .56** |
| WOMAC | .39* | -.22 | .40* | .37* | .17 | -.87** | .56** |  |

*Note.* PCS= Pain Catastrophising Scale, FFMQ= Five Factor Mindfulness Questionnaire, PSQI= Pittsburgh Sleep Quality Inventory, BDI= Becks Depression Inventory, STAI= State-Trait Anxiety Inventory, KOOS<sub>pain</sub>= Knee Injury and Osteoarthritis Score: Pain subscale, NRS= Numeric Rating Scale, WOMAC= Western Ontario and McMaster University Osteoarthritis Index. \* = $p < .05$ , \*\* = $p < .01$

### Pain Catastrophising and postsurgical outcomes

Pain catastrophising significantly predicted reductions in pain at 6-weeks (F(1,27)=4.53,p=.043,R^2^=.14,R^2^ =.11, SE_β= .41), 3-months (F(1,28)=12.1, p<.005, R^2^=.30,R^2^ =.28, SE_β= .35), 12-months (F(1,25)=4.61,p=.042, R^2^=.16,R^2^ =.12, SE_β= .43), and 24-months (F(1,17)=12.01,p<.005, R^2^=.41,R^2^ =.38, SE_β= .46). This suggests that patients who are high catastrophisers stand to gain the most beneficial reductions in pain following embolisation (Fig 4.).

**Figure 4.**
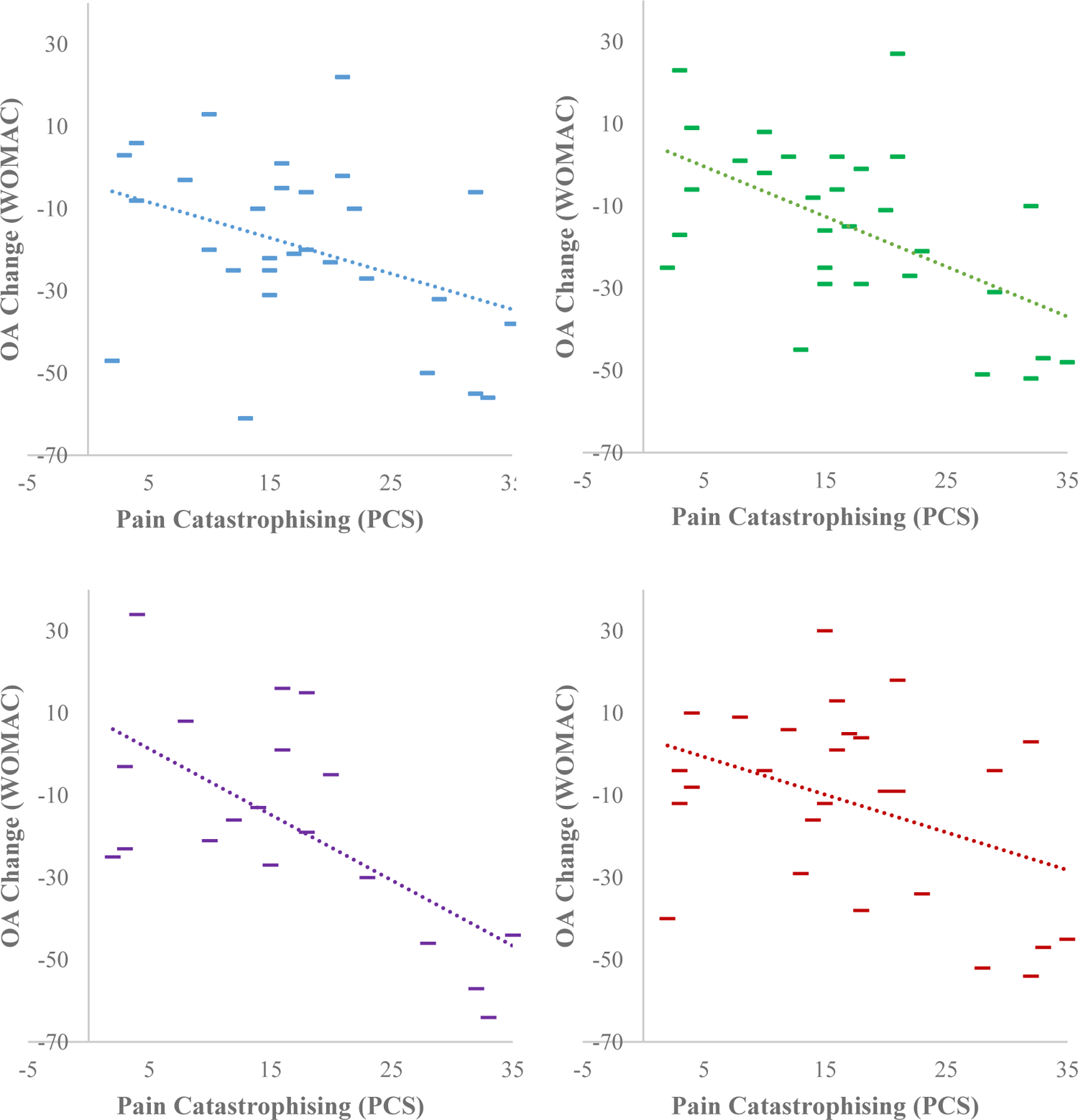
The association between baseline pain catastrophising, and reductions in baseline pain at 6-weeks, 3-months, 12-months and 24-months(clockwise). | Lower values represent decrease in WOMAC score over time, and improvement regarding OA impact)

### Association of Pain Catastrophising and functional connectivity of the dorsolateral frontal cortex

Analysis of rs-fMRI data revealed that patients with higher PCS scores were associated with higher connectivity between the dorsolateral prefrontal cortices and the somatosensory, motor, premotor, insula, operculum, anterior cingulate cortices (Fig.5; table 2).

**Figure 5.**
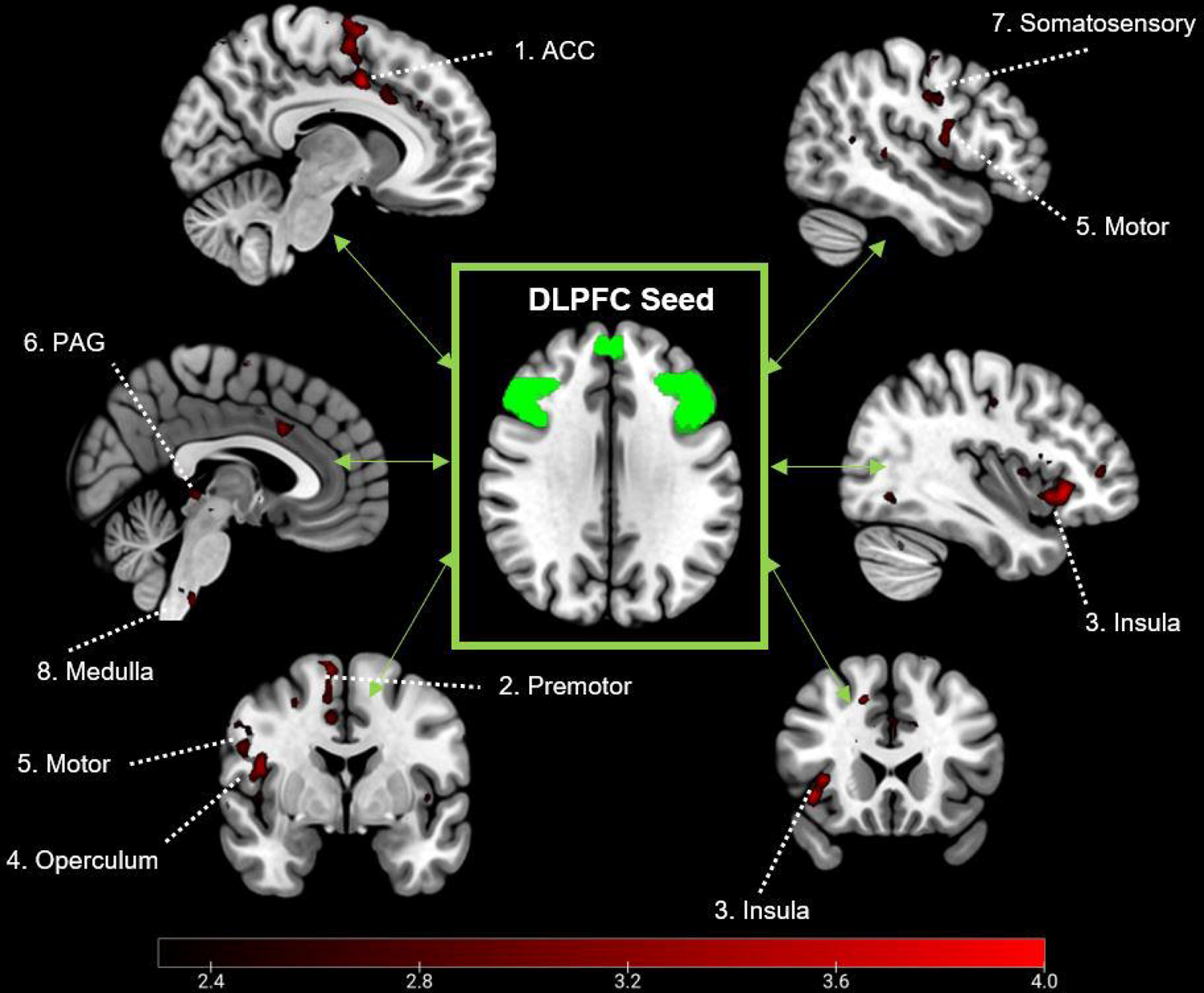
Clusters associated with connectivity to the DLPFC and pain catastrophising in MNI space (z<2.3) | Figure numbers relate to co-ordinates described in Table 2.

**Table 2.**
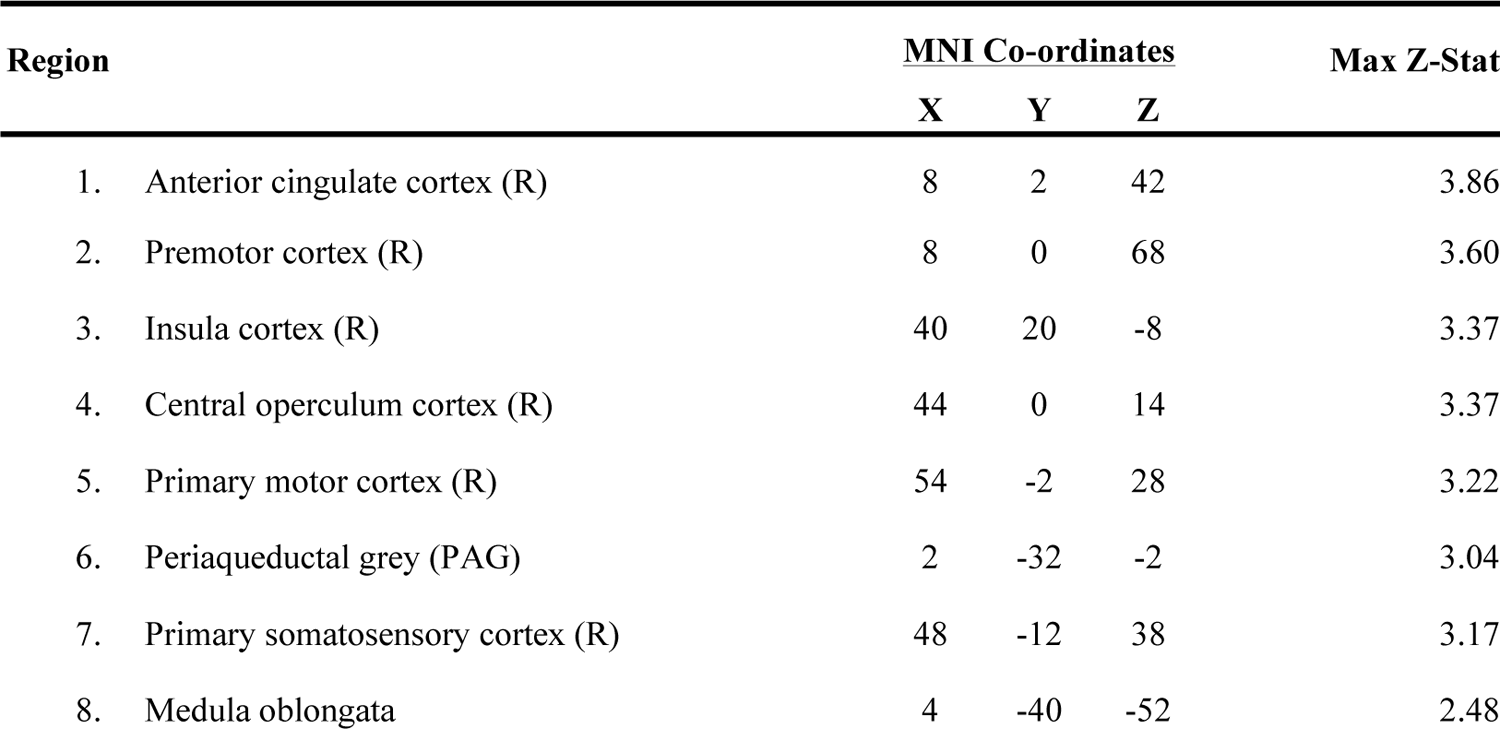
Statistical peaks of clusters in MNI space associated with pain catastrophising and functional connectivity to the DLPFC.

## Discussion

The study investigated if pain catastrophising can be used to predict clinical outcomes following genicular embolisation for the treatment of mild-moderate osteoarthritis. We also investigated how individual differences in catastrophising were associated with variations in functional connectivity of the DLPFC, a key pain modulatory region frequently associated with the underlying mechanisms of pain catastrophising[19,35,46,48]. We observed that, on average, patients experienced lasting reductions in pain as a result of the procedure, but that unexpectedly, those who were high catastrophisers at baseline gained the most profound improvements at all time-points (6-weeks, 3-months, 12-months and 24-months). Further, individual differences in PCS scores at baseline were associated with worse pain and a myriad of negative psychological impacts at baseline, as well as variations in functional connectivity at rest. We showed that pain catastrophising was associated with higher connectivity between the DLPFC and the anterior cingulate, premotor, motor, insula, operculum and somatosensory cortices.

Pain catastrophising is described as a set of maladaptive cognitions characterised by heightened pain intensity and unpleasantness[52,53], as well as an inability to disengage from the experience of pain[12]. Interestingly, our data indicate that high catastrophisers gained the most substantial reductions in pain following embolisation. For catastrophisers, pain represents an irrepressible aversive influence which cannot be disengaged from, as supported by our finding that PCS correlates with IAP scores. At baseline, catastrophising was associated with higher osteoarthritis pain, as well as associative negative lifestyle and psychological influences, such as depression, anxiety and poor sleep. The significant postsurgical decreases in pain may be most beneficial for those who find the impact of pain to be more intrusive, intense and unpleasant.

Interestingly, catastrophising has previously been linked to poor long-term outcomes following surgical intervention for osteoarthritic pain[21,43]. An important distinction between this experiment and similar previous studies, is that this study comprised solely of patients with mild-moderate osteoarthritis. Catastrophising has previously been shown to predict poor outcomes to invasive surgical procedures for knee osteoarthritis such as arthroplasty, often reserved for older patients with more severe or debilitating osteoarthritis [13], who are especially vulnerable to catastrophising[60]. Despite this, multiple studies have reported no association and challenged this position, stating the PC may be less trait-like and robust than initially thought[9,25]. One study investigating PC and recovery following total (TKA) and unicompartmental (UKA) joint replacement in 615 patients reported a similar finding to this study that high catastrophisers were associated with greater improvements on the Oxford Knee Score (OKS) scale[6]. Despite the lack of clarity on the direct influence of PC on postsurgical OA pain, it is logical that patients who have suffered for a longer duration from more intense and frequent pain may be at risk of more robust and entrenched pain and negative affect, whereas earlier treatment of mild-moderate OA stands to have more chance of malleable and impactful benefit.

Previous studies have also identified a moderating effect of treatment efficacy on the relationship between pain catastrophising and post-surgical pain[56]. Therefore, it may be that the beneficial outcomes for catastrophisers in this sample are because the successful treatment of their knee had a bifold impact on reductions in pain and pain catastrophising, as well. Pain catastrophising is often described as a robust, cognitive bias, representing a stable individual difference[52], although this position has been challenged more recently. It has been proposed that catastrophising may be a dynamic construct related to pain intensity[55], supported by its high malleability across varying interventions for surgical patients[18]. Successful clinical treatment may theoretically alter responses to multiple items in the PCS such as “It’s terrible and I think it’s never going to get any better” (item 3) or “there’s nothing I can do to reduce the intensity of pain” (item 12)[51]. In these instances, the entrenchment of pain may not yet have taken hold, and maladaptive cognitive biases can still be challenged via successful alleviation of pain. However, as this study only collected PCS scores at baseline, future studies should investigate the impact of successful clinical intervention on the stability of PCS scores for patients with non-severe OA.

Alongside the behavioural findings of PCS, this study also provides insight to the neural mechanisms underlying individual differences in catastrophising. Patients completed rs-fMRI scans, where no task was administered, to examine intrinsic functional connectivity. Evidence has frequently identified the DLPFC as a key region in the process of catastrophising, associated with the interpretation of pain and descending pain modulation[46–48]. Our data indicates that catastrophising is associated with higher connectivity between the DLPFC, and multiple areas of the brain associated with sensory-discriminative processing of pain (motor, sensorimotor, premotor cortices), attentive-perception and salience of pain (insula and anterior cingulate cortices) and modulation of pain encoded within the brainstem (medulla and PAG).

The interpretation of rs-fMRI findings must be evaluated conservatively, as many direct explanations of pain processing would require event-related stimulation to empirically test. Additionally, this finding requires replication due to the relatively low sample size[20], and this follow-up will be completed in the second phase of the study. However, as catastrophising facilitates heightened salience and attentional focus towards pain, increased functional connectivity of the DLPFC with regions involved in processing of pain may represent an increased demand for endogenous modulation. These findings complement previous work by Seminowicz and Davis[46], who found that during mild painful stimulation, PCS was associated with increases in activity in the DLPFC, as well as in the insula, motor and rostral anterior cingulate cortices, matching clusters identified from our analysis. Conversely, if the intensity of pain is increased, PCS was then associated with decreased activity in the DLPFC. The authors proposed that during intense pain, high catastrophisers may have difficulty disengaging from pain, via a lack of top-down control.

This dynamic relationship between PCS and pain intensity may provide an explanation as to why PCS is associated with poor outcomes in severe OA, whereas when mild-moderate OA is treated successfully via embolisation, these patients experience a reduction in pain which they can then continue to effectively modulate.

The current study is concordant with the interim analysis by Little et al[34]., suggesting that genicular arterial embolisation is an effective treatment for mild-moderate osteoarthritis pain. Pre-surgically, pain catastrophising is associated with a range of negative co-morbidities, such as depression, anxiety and increased pain. However, our data suggests that patients who are high catastrophisers stand to gain the most benefit from successful intervention, and the PCS can predict outcomes up to 12-months post-surgically. Neural data suggests that catastrophisers have a higher functional integration of descending modulatory and pain processing regions in the brain, and that the DLPFC is a key region in this process. These results suggest that the increased attentional preoccupation with pain at baseline, facilitates a heightened requirement for pain modulation, which is supported by previous studies[46]. A follow-up trial will aim to replicate these findings within a larger sample in a randomly controlled trial (RCT), alongside sham surgery, to evaluate embolisation alongside a suitable control.

## Data Availability

All data produced in the present study are available upon reasonable request to the authors

---

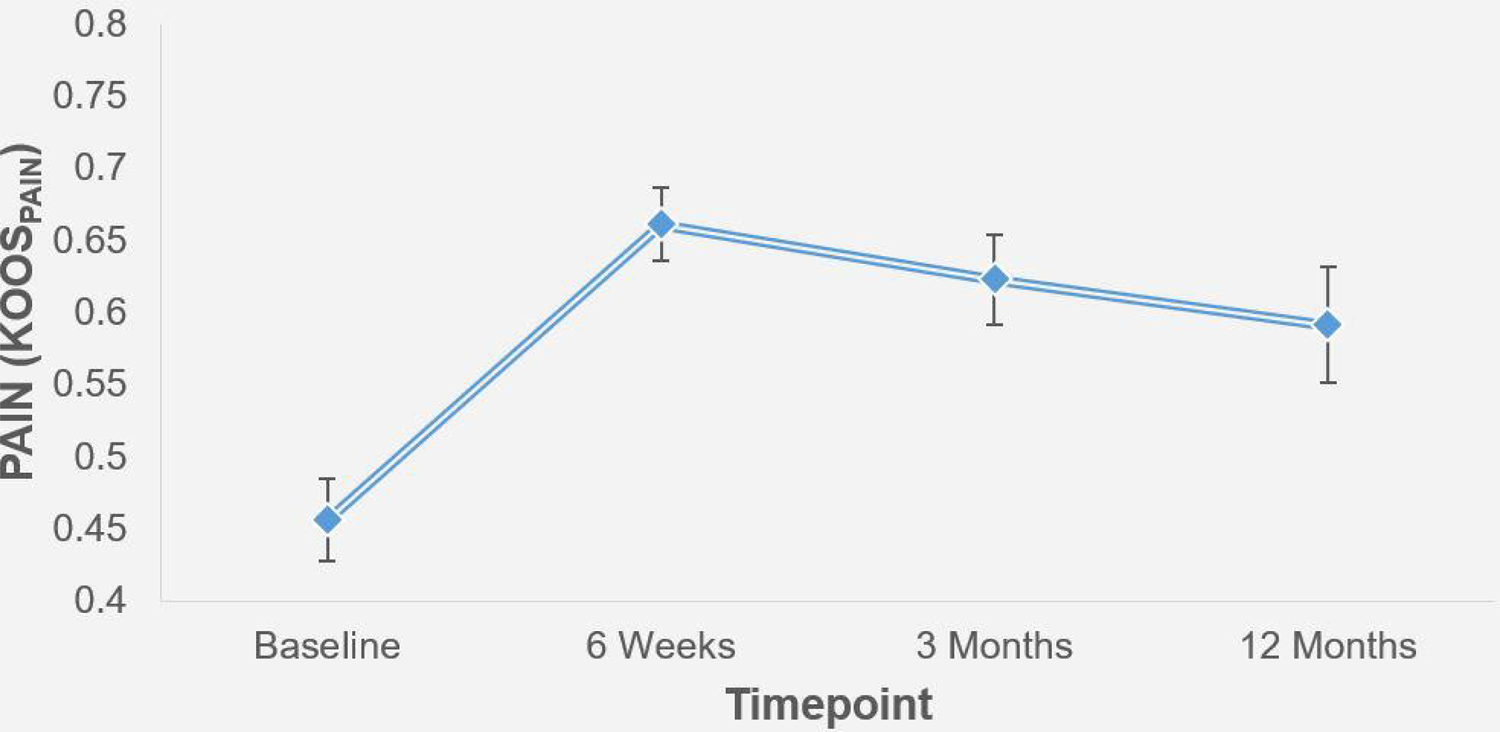

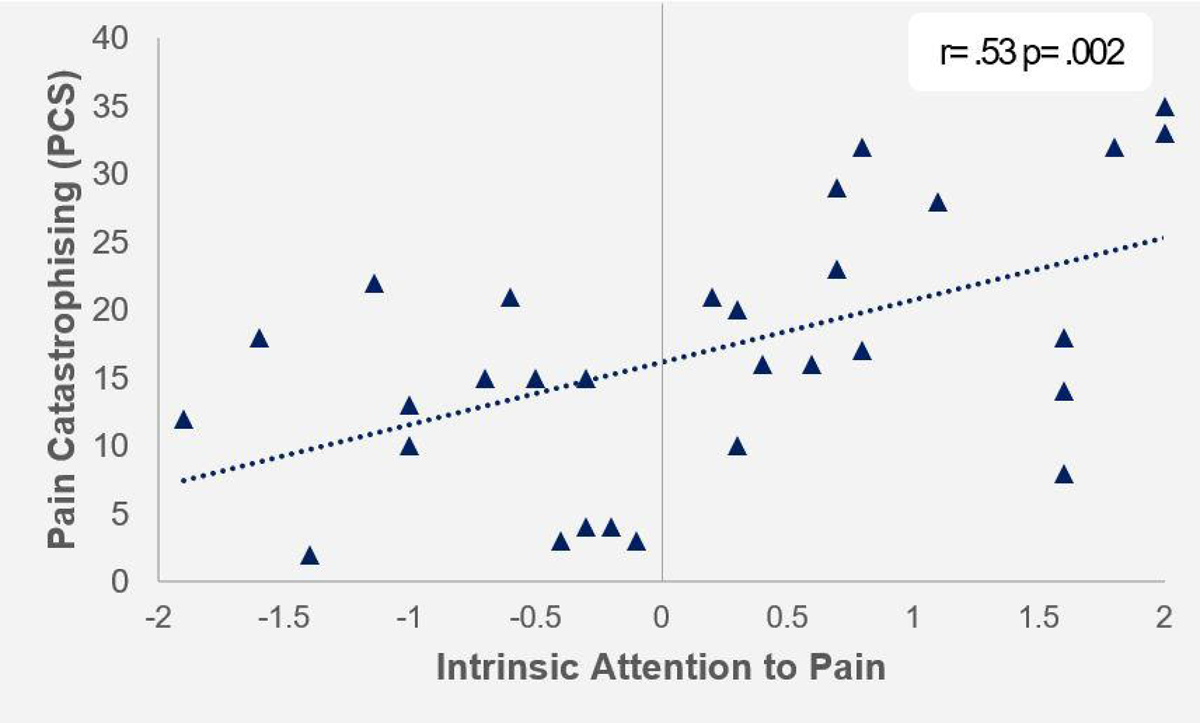

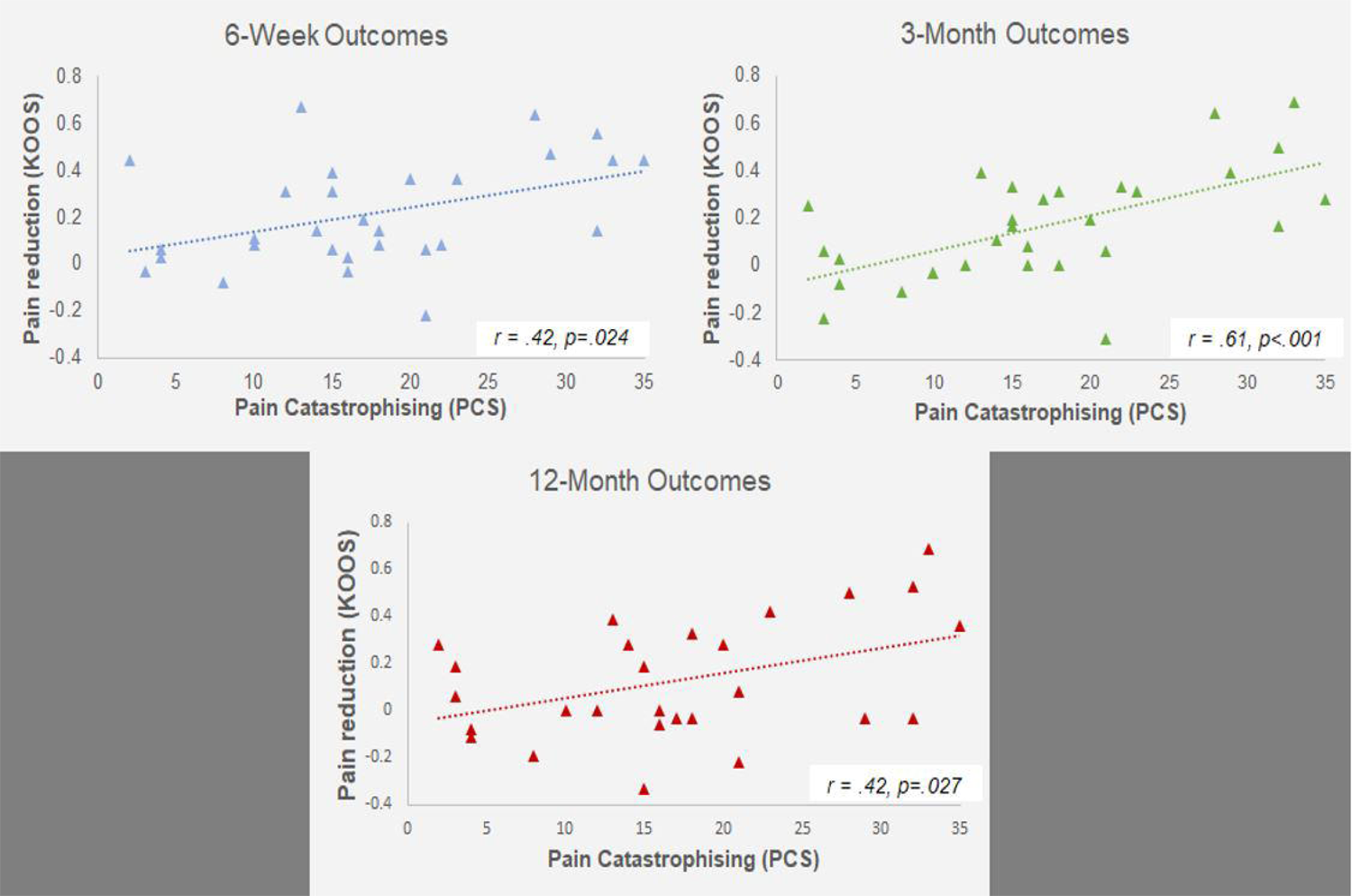

